# Consistent Performance of GPT-4o in Rare Disease Diagnosis Across Nine Languages and 4967 Cases

**DOI:** 10.1101/2025.02.26.25322769

**Authors:** Leonardo Chimirri, J. Harry Caufield, Yasemin Bridges, Nicolas Matentzoglu, Michael Gargano, Mario Cazalla, Shihan Chen, Daniel Danis, Alexander JM Dingemans, Petra Gehle, Adam S.L. Graefe, Weihong Gu, Markus S. Ladewig, Pablo Lapunzina, Julián Nevado, Enock Niyonkuru, Soichi Ogishima, Dominik Seelow, Jair A Tenorio Castaño, Marek Turnovec, Bert BA de Vries, Kai Wang, Kyran Wissink, Zafer Yüksel, Gabriele Zucca, Melissa A. Haendel, Christopher J. Mungall, Justin Reese, Peter N. Robinson

**Affiliations:** Berlin Institute of Health at Charité – Universitätsmedizin Berlin, Berlin, Germany; Lawrence Berkeley National Laboratory, Berkeley, CA, USA; William Harvey Research Institute, Barts and The London School of Medicine and Dentistry, Queen Mary University of London, London, UK; Semanticly, Athens, Greece; The Jackson Laboratory for Genomic Medicine; INGEMM-Idipaz, Institute of Medical and Molecular Genetics, Hospital Universitario La Paz, Madrid, Spain; Department of Pathology and Laboratory Medicine, University of Pennsylvania, Philadelphia, PA, USA; Department of Human Genetics, Donders Institute for Brain, Cognition and Behaviour, Radboud University Medical Center, Nijmegen, the Netherlands; Deutsches Herzzentrum der Charité, Berlin, Germany; Chinese HPO Consortium, Beijing, China; Department of Ophthalmology, University Clinic Marburg - Campus Fulda, Fulda, Germany CIBERER-ISCIII, Madrid, Spain; Trinity College, Hartford, CT, USA; INGEM/ToMMo, Tohoku University, Miyagi, Japan; Department of Biology and Medical Genetics, 2nd Faculty of Medicine, Charles University in Prague and Motol University Hospital, Prague, Czech Republic; Utrecht University, Utrecht, Netherlands; Department of Human Genetics, Bioscientia Healthcare GmbH, Ingelheim, Germany; Institute for Maternal and Child Health - IRCCS “Burlo Garofolo” - Trieste, Trieste 34137, Italy; University of North Carolina at Chapel Hill

## Abstract

**Background:** Large language models (LLMs) are increasingly used in the medical field for diverse applications including differential diagnostic support. The estimated training data used to create LLMs such as the Generative Pretrained Transformer (GPT) predominantly consist of English-language texts, but LLMs could be used across the globe to support diagnostics if language barriers could be overcome. Initial pilot studies on the utility of LLMs for differential diagnosis in languages other than English have shown promise, but a large-scale assessment on the relative performance of these models in a variety of European and non-European languages on a comprehensive corpus of challenging rare-disease cases is lacking.

**Methods:** We created 4967 clinical vignettes using structured data captured with Human Phenotype Ontology (HPO) terms with the Global Alliance for Genomics and Health (GA4GH) Phenopacket Schema. These clinical vignettes span a total of 378 distinct genetic diseases with 2618 associated phenotypic features. We used translations of the Human Phenotype Ontology together with language-specific templates to generate prompts in English, Chinese, Czech, Dutch, German, Italian, Japanese, Spanish, and Turkish. We applied GPT-4o, version gpt-4o-2024-08-06, to the task of delivering a ranked differential diagnosis using a zero-shot prompt. An ontology-based approach with the Mondo disease ontology was used to map synonyms and to map disease subtypes to clinical diagnoses in order to automate evaluation of LLM responses.

**Findings:** For English, GPT-4o placed the correct diagnosis at the first rank 19·8% and within the top-3 ranks 27·0% of the time. In comparison, for the eight non-English languages tested here the correct diagnosis was placed at rank 1 between 16·9% and 20·5%, within top-3 between 25·3% and 27·7% of cases.

**Interpretation:** The differential diagnostic performance of GPT-4o across a comprehensive corpus of rare-disease cases was consistent across the nine languages tested. This suggests that LLMs such as GPT-4o may have utility in non-English clinical settings.

**Funding:** NHGRI 5U24HG011449 and 5RM1HG010860. P.N.R. was supported by a Professorship of the Alexander von Humboldt Foundation; P.L. was supported by a National Grant (PMP21/00063 ONTOPREC-ISCIII, Fondos FEDER).

## Introduction

Large language models (LLM) are being carefully investigated in the medical domain owing to their linguistic and problem solving capabilities. These artificial intelligence (AI) models are pre-trained by ingesting large amounts of unlabeled data in order to learn to generate coherent text, thereby opening up an array of possibilities in disparate aspects of clinical practice. Moreover, LLMs have been shown to encode latent medical knowledge and to be able to carry out deductive reasoning, which would make them candidate assistance tools in hard-to-diagnose, complex clinical cases.^1^

The majority of medical literature and therefore the LLMs’ relevant training data is in English. According to CommonCrawl,^2^ an open repository of web crawl data that can be used to estimate the distribution of internet data used by LLMs for training, 43% of available web pages are in English (version CC-MAIN-2024-51). The percentages for the other eight languages in our study range from 1·0% (Czech) to 5·4% (German). While the precise training data for most LLMs are not publicly disclosed, estimates suggest a high proportion of English language content.^3^ Thus, more material is available for training in English than in any other language, which may imply an expectation of LLMs to achieve higher performance in English, as suggested from previous studies.^4–6^ This clearly would have implications for their integration in clinical practice, as well as on the topic of AI fairness in the health domain. Few studies about multilinguality of generalist LLMs in biomedicine have been carried out, most of which tested the performance of a LLM on medical licensing exams or standardized medical questions in one non-English language or one language as compared to English (see Japanese,^7^ in-context enhanced Chinese,^8^ German,^9^ and Arabic^6^).

Roughly 25% of rare disease patients go 5-30 years without a diagnosis, and 40% of initial diagnoses are wrong.^10^ There are at least 10,000 RDs,^11^ and the diagnostic yield of genomic sequencing is still low (25– 50%).^12^ Therefore, the differential diagnosis of RDs represents a challenging task with which to evaluate the capabilities of LLMs.

LLMs have shown promise in supporting the differential diagnosis of RDs in English,^13^ yet most of the earth’s population does not use English as their first language and clinical practice is carried out in diverse languages across the world. In this paper, we assess the relative performance of LLMs in RD differential diagnostics by creating a large multilingual set of published patient descriptions. Thereby we address limitations of previous studies that used simulated/synthetic patients or small cohorts and provide a comprehensive analysis of the relative performance of a leading LLM in nine different languages.

## Methods

### Overview of Study

We conducted a comparison of GPT-4o’s ability in genetic differential diagnostics across several languages. We analyzed 4967 case reports from the literature and generated LLM prompts in nine languages and directed GPT-4o to return a ranked list of possible diagnoses for each case. The Mondo disease ontology,^14^ which contains numerous synonyms for each disease, was used to map the diagnoses returned by GPT-4o to a unique standardized medical vocabulary, to which we applied our automatic ontology-aware scoring.^15^ The rank of the correct diagnosis in the output was compared across languages in order to assess to what degree the LLM’s performance is prompt-language dependent. This study is reported according to the TRIPOD-LLM reporting guideline.^16^

### Human Phenotype Ontology Internationalization

The Human Phenotype Ontology (HPO) provides a standardized vocabulary of 19,034 terms that describe the phenotypic abnormalities of human disease. Version 2024-12-12 was used for this study. Additionally, the HPO provides a comprehensive corpus of phenotype annotations (HPOA) that form computational models of 8333 rare diseases. HPO applications include genomic interpretation for diagnostics, gene-disease discovery, machine learning (ML) and electronic health record (EHR) cohort analytics.^17^ The HPO Internationalization Effort comprises language-specific working groups that have translated HPO term labels and in some cases synonyms and definitions from English into other languages.^18^ For the current project, we used the Chinese, Czech, Dutch, German, Italian, Japanese, Spanish, and Turkish translations. All translations are freely available (see data availability section). Information about the number of available translated terms present in HPO is found in supplemental table S1. Translations are created by or confirmed by human experts before inclusion in a HPO release.

### Structured Data from Case Reports: Phenopackets

The Global Alliance for Genomics and Health (GA4GH) Phenopacket Schema is a standard for sharing phenotypic, genetic and clinical information.^19^ The Phenopacket Schema obtained International Standard Organization approval as ISO 4454:2022. Each Phenopacket is a clinical vignette about one individual with representations of phenotypic abnormalities using HPO terms, as well as a specification of the disease diagnosis and other information.

The Phenopackets used in this project were selected from the Phenopacket Store version 0.1.19, an openly available collection of Phenopackets manually curated from published case reports.^20^ It contains a total of 6668 Phenopackets representing 475 diseases and 423 disease genes. We restricted our analysis to the subset of Phenopackets in the Phenopacket Store for which translations of all associated HPO terms exist in the eight languages mentioned above. This yielded a dataset of 4967 Phenopackets from 729 PubMed IDs, comprising 343 causative disease genes, 378 diseases, 2934 alleles, 2618 unique HPO terms and an average of 14 HPO terms per patient.

### Prompt generation

Phenopackets comprise a hierarchical structure that is typically stored as a JSON file. We developed a strategy to create narrative prompts from each Phenopacket by a templating system implemented in a Java application called phenopacket2prompt.^21^ Each template consists of constant texts (such as the header that instructs GPT to return a differential diagnosis), and a series of templates to represent the age and sex of the individual represented by the Phenopacket as well as phenotypic abnormalities that were observed or excluded. If available, the age of onset of the disease or specific manifestations is recorded. The templating system involves vocabulary for describing the individual in each of the languages. HPO terms in each of the languages are substituted into the corresponding templates. The translators of each of the eight languages are physicians or medical researchers, and an example of the part of a prompt describing a patient is shown in figure 1. The correctness of the translation templates was confirmed by review of 54 simulated cases that were output in each language using permutations of ages, sexes, as well as observed and excluded HPO terms. The diagnosis and the genetic information were not included in the prompts. At the end of the constant section, we state that the case is a genetic disease and we request GPT to return an ordered list of candidate diagnoses, giving an example output. The example output is always given in English and in non-English languages we explicitly instruct GPT to return the differential diagnosis in English. An example Phenopacket with associated full prompt in English and its translation into the eight languages is available in supplementary tables S3-S4.

**Figure 1.**
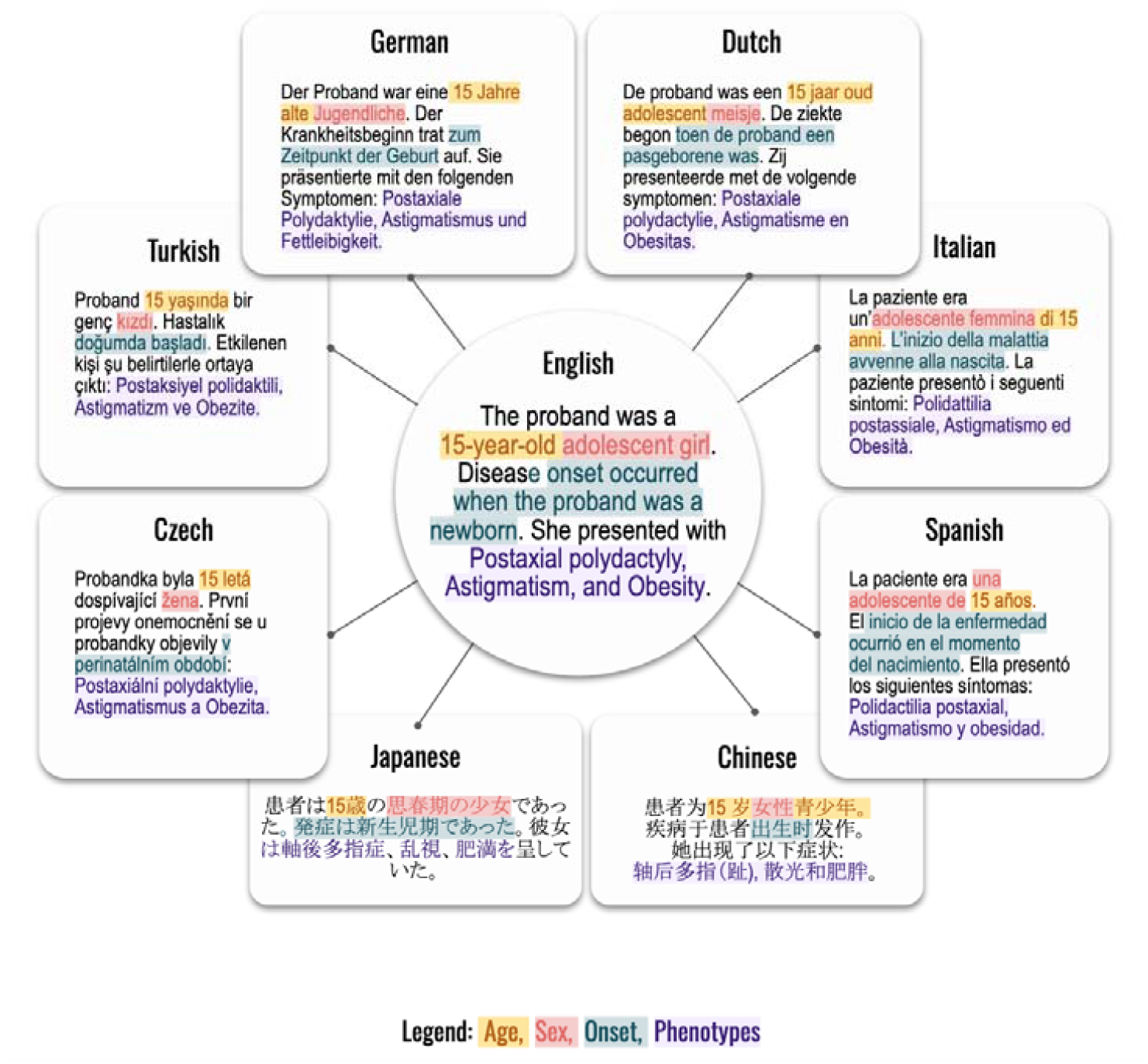
Templated system for generating prompts using translation of the HPO into 8 languages. An excerpt of one prompt is shown. Shading indicates Age, Sex, Onset, HPO Phenotypes.

### Grounding and Scoring

The queries to GPT-4o were carried out through its API between November 22nd, 2024 and January 20th, 2025. The knowledge cutoff claimed by openAI is October 2023 for all GPT-4o models, including gpt-4o-2024-08-06, the version we used. We used the default parameters, with no token length limitation and using unit temperature. We instructed the LLM to reply in the form of free text (e.g., Marfan syndrome), rather than a corresponding ontology term identifier (e.g., MONDO:0007947), because the task of returning identifiers may be prone to so-called “hallucinations”, where plausible-sounding yet wrong answers are given.^22,23^ We leveraged our pheval.llm pipeline to parse the LLM response for free-text candidate diagnoses, to identify the corresponding ontology identifier, and to rank genetic subforms of a clinical disease as equivalent (e.g., Loeys-Dietz syndrome and any of its six genetic subforms were regarded as equivalent for the purposes of assessing correctness of the differential diagnosis). PhEval.llm is a freely available plugin for the PhEval framework and leverages Monarch Initiative LLM tooling.^15,24,25^

With this, we could computationally score the responses of GPT for all Phenopackets in all nine languages, corresponding to 44,198 differential diagnoses with more than 210 thousand candidate diseases (of which 6733 unique guesses by GPT). Finally, we computed the number of correct diagnoses found at the top of the differential diagnosis (“Top-1”), within the first three candidate diseases (“Top-3”), and likewise for “Top-10”. This procedure was carried out for all nine languages.

### Role of the funding source

The funding sources had no role in study design, data collection, data analysis, data interpretation, writing of the report, and decision to submit.

## Results

In this work we leveraged translations of the Human Phenotype Ontology into Chinese, Czech, Dutch, German, Italian, Japanese, Spanish, and Turkish to test the relative performance of GPT-4o in differential diagnostic support. To do so, we leveraged GA4GH Phenopackets, representing the clinical data (phenotypic abnormalities and diagnosis) for 4967 patients with 378 diseases drawn from 729 publications. We generated a programmatic templating system that generated a narrative text using templates to represent the age, sex, and age of onset of disease together with observed and excluded phenotypic features (figure 1).

Results are counts of correct diagnoses at a given rank in the differential diagnosis, shown in table 1 aggregated at “Top-1”, “Top-3”, and “Top-10”. The “Not ranked” column shows the number of cases in which the reply of GPT-4o did not contain the correct diagnosis. Additionally, the “No Diagnosis” column indicates the number of cases in which GPT did not return text that could be parsed as a differential (for example, “I’m sorry but based on the information provided, I cannot return a confident diagnosis”). In no case did we observe a correct result beyond rank 10, so that the sum of the last three columns in table 1 is always 4967, the total number of cases. Depending on the language, between 2·3% and 5·6% items in the differential could not be successfully grounded (i.e., the Mondo term corresponding to the diagnosis could not be identified; see supplemental table S2).

**Table 1:** Results of differential diagnosis by GPT-4o by language. Numbers indicate counts, not ranked includes grounding failures but not instances of refusal by GPT to deliver a diagnosis, which is counted separately in the rightmost column “No Diagnosis”.

| Language | Top-1 | Top-3 | Top-10 | Not Ranked | No Diagnosis |
| --- | --- | --- | --- | --- | --- |
| English | 985 | 1340 | 1546 | 3420 | 1 |
| Chinese | 917 | 1352 | 1417 | 3544 | 6 |
| Czech | 887 | 1252 | 1409 | 3532 | 26 |
| Dutch | 1013 | 1370 | 1521 | 3423 | 23 |
| German | 944 | 1304 | 1455 | 3411 | 101 |
| Italian | 860 | 1327 | 1514 | 3450 | 3 |
| Japanese | 796 | 1256 | 1364 | 3348 | 255 |
| Spanish | 951 | 1370 | 1579 | 3388 | 0 |
| Turkish | 903 | 1311 | 1449 | 3473 | 45 |

In figure 2 we show frequencies obtained as Top-N divided by the total number of cases excluding those in “No Diagnosis”. To test for statistical differences between ranks in different languages we used SciPy’s ‘stats’ package to perform a Kruskal-Wallis^26^ H-test obtaining as a statistic H=22·3 and p-value=0·0044, indicating statistically significant differences between the languages. English was the second best performer at Top-1,Top-3, and Top-10. At Top-1, there were 19·8% of correct cases in English while other languages ranged from 16·9% to 20·5%. At Top-3, the spread decreases, with the correct diagnosis among the Top-3 candidates in the differential diagnosis 27·0% of cases for English, compared to the range 25·3% to 27·7% for the other languages. This comprises a relative difference of *at most* 6% for Top-3 with respect to English, and at most 8% for Top-10, where English scored 31·1% while other languages lie in the range 28·5% to 31·8%.

**Figure 2.**
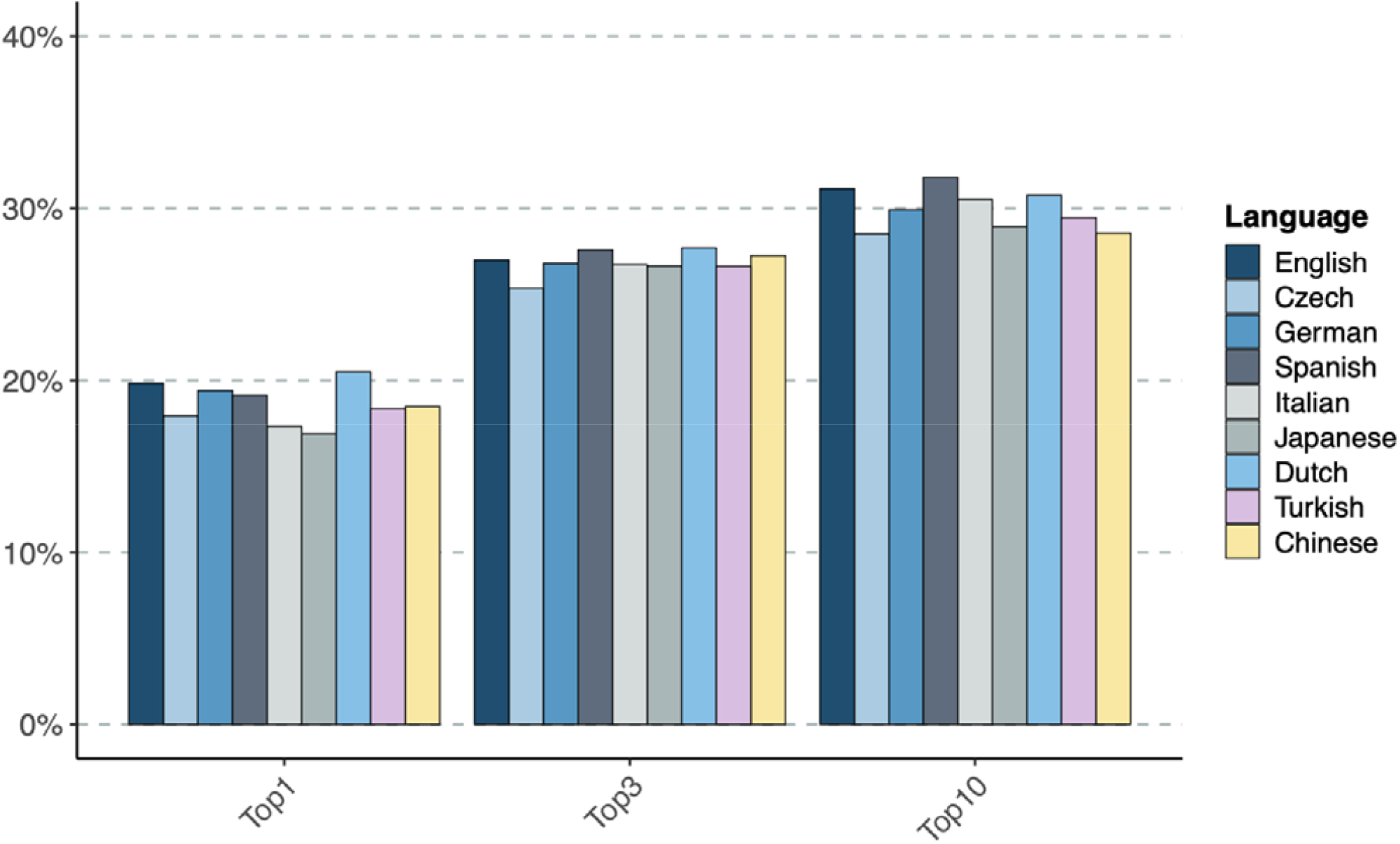
Differential diagnostic performance of GPT-4o in English, Chinese, Czech, Dutch, German, Italian, Japanese, Spanish, and Turkish. The percentage of cases in which GPT-4o place the correct diagnosis in rank 1 (Top-1), within the top three ranks (Top-3) or within the first ten ranks (Top-10) is shown.

## Discussion

We prompted the GPT-4o language model with 4967 RD cases in nine different languages. All languages in this study constitute at least ∼1% of the CommonCrawl, which is a proxy for the amount of relative internet data available in a given language, a reflection of the language-specific data available for training. For these nine languages we have shown that GPT-4o is able to perform differential diagnostics for RD with nearly the same performance. This would be surprising if LLMs such as GPT only used language-specific models to answer queries because most of its training data and of relevant medical literature is in English. Our result suggests that this model can generalize medically relevant knowledge derived mainly from English language texts in order to answer queries posed in (at least) the eight non-English languages tested.

The creation of a large set of realistic vignette-like prompts originating from real cases and translated in multiple languages overcomes limitations of previous diagnostics studies with LLMs, such as the utilization of extensive and unrealistically long case reports,^27^ the usage of simulated/synthetic patients,^28,29^ the usage of comparably small cohorts,^27–29^ and the difference in style and length of real clinical notes coming from different countries.^29^

Our study has several limitations. We used the same zero-shot prompting strategy for each language and did not attempt to improve performance using more sophisticated strategies such as chain of thought or retrieval augmented generation approaches.^30^ Our evaluation made use of lists of phenotype terms, rather than narrative clinical notes, and thus may not reflect challenges or nuances relating to individual languages. Additionally, we only tested a specific GPT model, and were only able to test a selection of relatively widely used European and Asian languages. Therefore, further research will be needed to determine the relative performance of LLMs on a wider range of languages using more prompting strategies. In order to be able to automatically evaluate the response of GPT, we explicitly asked for the LLM replies to be in English. We are not able to say if failures in mapping free text to Mondo identifiers observed in this step were responsible for the slightly worse performance of some languages in our testing. We did not assess the behavior and accuracy for multilingual diagnostics of other LLMs in our study, but our results with GPT-4o suggest that in principle it is possible to train an LLM to leverage medical knowledge that is predominantly recorded in English to support differential diagnostics in other languages.

GPT-4o’s ability to carry out diagnostics of complex cases in different languages is notable considering it is a general purpose language model predominantly trained with English data. OpenAI claims to have significantly increased the multilingual capabilities of GPT-4o with respect to its previous models.^31^

Consistent performance across languages has implications for the implementation of these models in clinical practice across the globe. Many people in low- and middle-income countries (LMICs) have limited access to healthcare services.^32^ As LLMs become increasingly proficient in supporting differential diagnosis and related domains such as bedside consultation question answering and addressing questions from the general public,^33^ there is a great potential to improve care for people in LMICs by supplementing existing systems with LLM-driven services. It would be desirable to offer such services in local languages, especially for consumer-facing applications. Future work will be required to assess performance of LLMs in LMICs (all languages assessed in our study are from high-income countries).

## Data availability

HPO translations: https://obophenotype.github.io/hpo-translations/

4967 phenopackets and corresponding prompts in English, Chinese, Czech, Dutch, German, Italian, Japanese, Spanish, and Turkish, together with all our results at DOI: https://zenodo.org/records/14907503 All code is publicly available at:

pheval.llm: https://github.com/monarch-initiative/pheval.llm

ontology access kit: https://github.com/INCATools/ontology-access-kit

phenopacket2prompt: https://github.com/monarch-initiative/phenopacket2prompt

## Contributors

L.C. and J.R. created the pheval.llm python code with input from J.H.C and Y.B. L.C. wrote the first draft of the paper in close collaboration with P.N.R. The phenopacket2prompt Java application was written by P.N.R. and L.C. who also wrote the Italian prompts and coordinated the language specific developers of the prompts. M.H. conceptualized the ontology translational strategy and Phenopackets. N.M. developed much of the technical infrastructure around translation management and is the main liaison for the HPO translation community.

P.N.R. and J.R. led the overall study. C.M. devised the Mondo evaluation approach and advised on the use of LLMs.

C.S. coded the Chinese prompts; G.W. is part of the development of Chinese HPO; K.W. confirmed the Chinese translations; D.D. coded the Czech prompts; M.T. provided Czech translations and confirmations; A.S.L.G. coded the Turkish prompts and helped with Turkish translations; Z.Y. confirmed and provided Turkish translations; D.S. suggested and confirmed German translations; G.J.P. confirmed the German translations; G.Z. provided the Italian translations; J.N., J.A.T.C, M.C., and P.L. provided the Spanish translations; K.W. coded the Dutch prompts; A.j.M.D. and B.B.A.d.V. provided the Dutch translations; S.O. provided the Japanese translations. All authors had access to the manuscript and take responsibility for the decision to submit the publication. All of the data used in this study is publicly available.

## Declaration of interests

MH is a co-founder of Alamya Health.

## Acknowledgements

We acknowledge funding from NHGRI 5U24HG011449, 5RM1HG010860, and R24OD011883. P.N.R. was supported by a Professorship of the Alexander von Humboldt Foundation; P.L. was supported by a National Grant (PMP21/00063 ONTOPREC-ISCIII, Fondos FEDER). CM and JR were supported in part by the Director, Office of Science, Office of Basic Energy Sciences, of the US Department of Energy (Contract No. DE-AC0205CH11231).

## Notes

### Author Declarations

We analyzed 4967 published case reports from the literature taken from Phenopacket Store: Danis D, Bamshad MJ, Bridges Y, Caballero-Oteyza A, Cacheiro P, Carmody LC, Chimirri L, Chong JX, Coleman B, Dalgleish R, Freeman PJ, Graefe ASL, Groza T, Hansen P, Jacobsen JOB, Klocperk A, Kusters M, Ladewig MS, Marcello AJ, Mattina T, Mungall CJ, Munoz-Torres MC, Reese JT, Rehburg F, Reis BCS, Schuetz C, Smedley D, Strauss T, Sundaramurthi JC, Thun S, Wissink K, Wagstaff JF, Zocche D, Haendel MA, Robinson PN. A corpus of GA4GH phenopackets: Case-level phenotyping for genomic diagnostics and discovery. HGG Adv. 2025 Jan 9;6(1):100371. doi: 10.1016/j.xhgg.2024.100371. Epub 2024 Oct 10. PMID: 39394689; PMCID: PMC11564936.

