## Supplementary material for "Consistent Performance of GPT-4o in Rare Disease Diagnosis Across Nine Languages and 4967 Cases": Online supplement

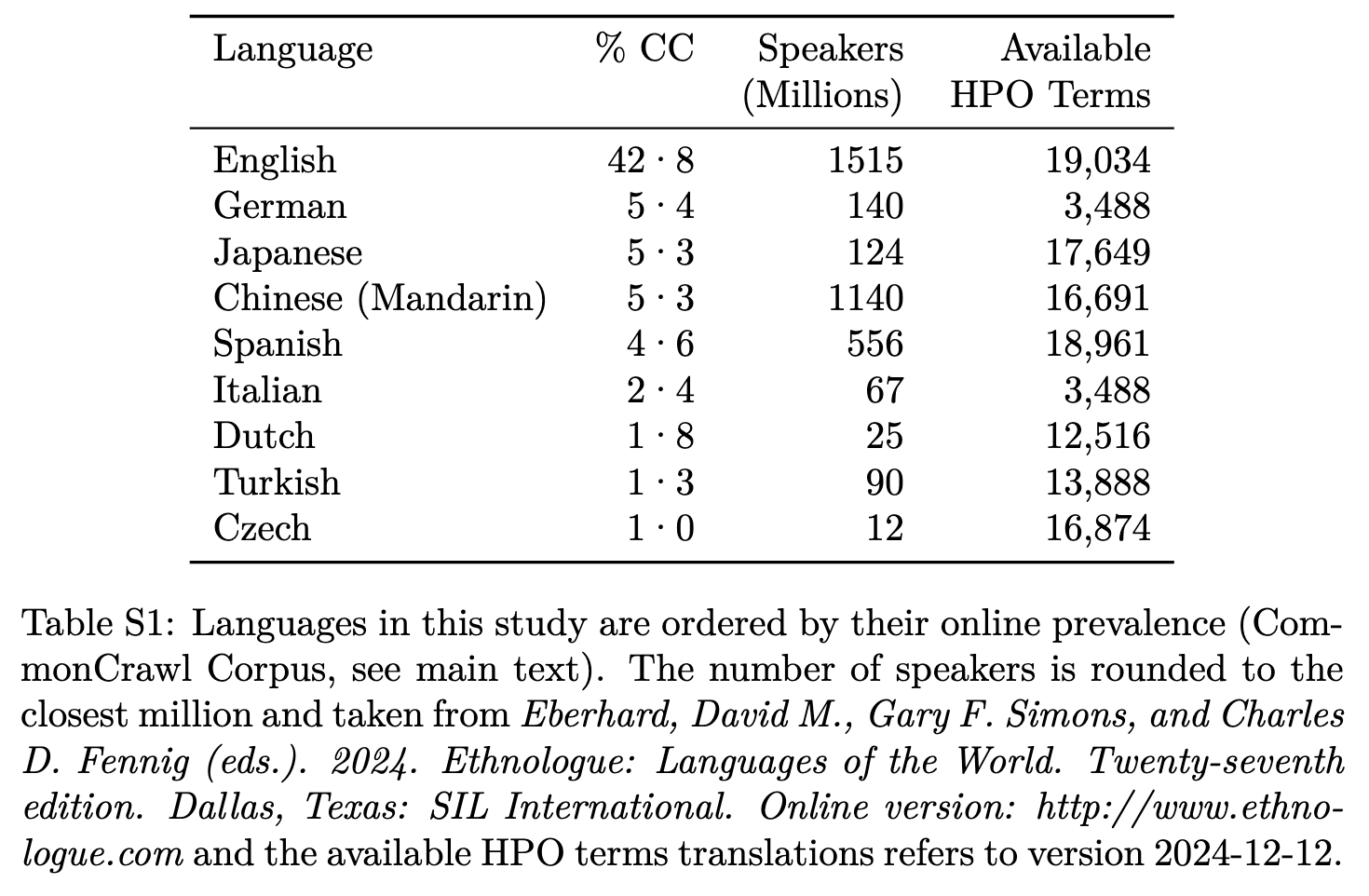

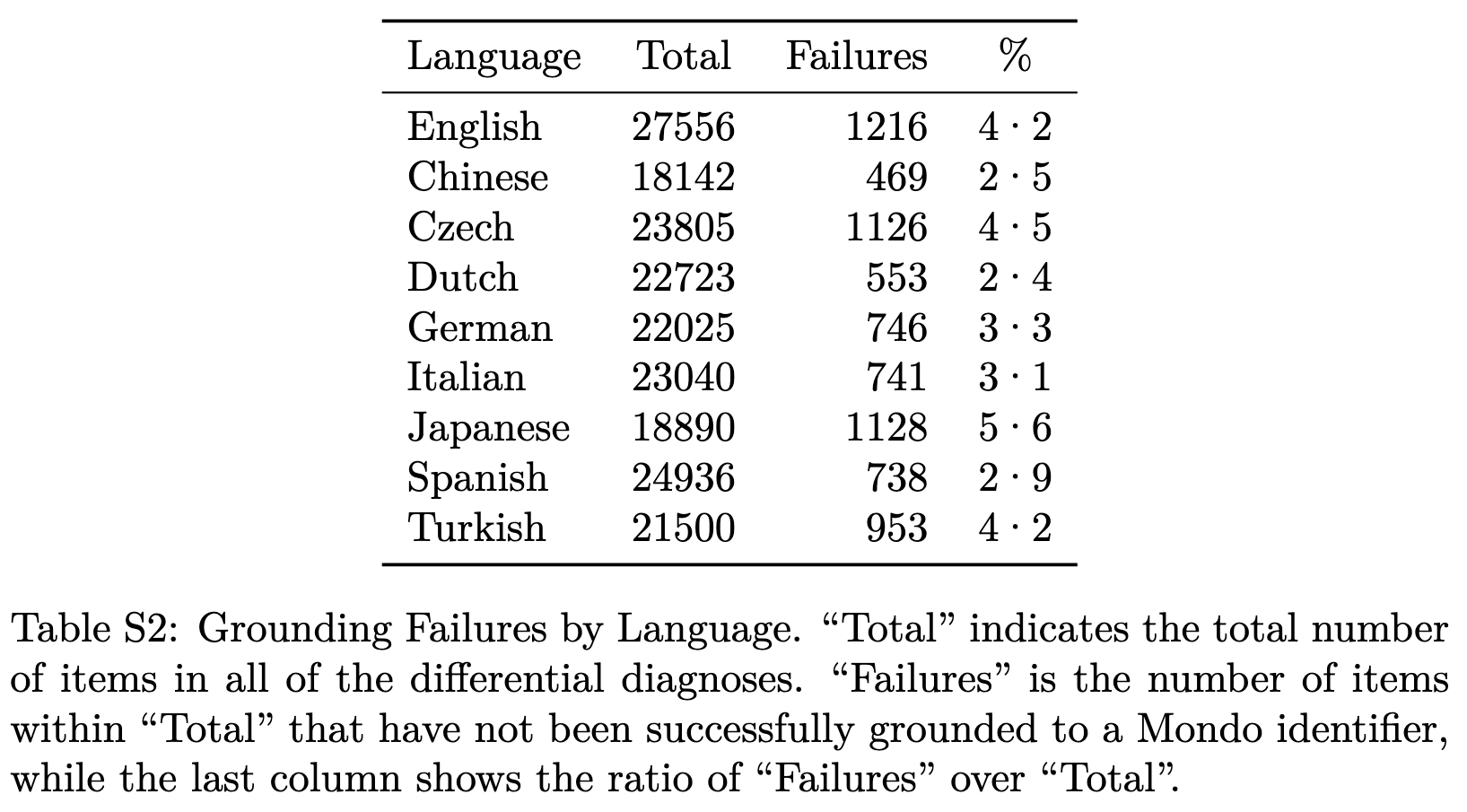

**Table S3.** An example phenopacket in JSON format and the corresponding full prompt in English.

Showing only parts of the phenopacket used in the study, the full one can be seen at https://github.com/monarch-initiative/phenopacket-store/blob/80a982726b4d8253a2230aa47089f371f9b5373b/notebooks/BBS1/phenopackets/PMID_22940089_PT2.json

| JSON | JSON (continued) | English Prompt |
| --- | --- | --- |
| {  "id": "PMID_22940089_PT2",  "subject": {  "id": "PT2",  "timeAtLastEncounter": {  "age": {  "iso8601duration": "P15Y"  }  },  "sex": "FEMALE"  },  "phenotypicFeatures": [  {  "type": {  "id": "HP:0007663",  "label": "Reduced visual acuity"  },  "onset": {  "age": {  "iso8601duration": "P3Y"  }  }  },  {  "type": {  "id": "HP:0000662",  "label": "Nyctalopia"  },  "onset": {  "age": {  "iso8601duration": "P7Y"  }  }  },  {  "type": {  "id": "HP:0100259",  "label": "Postaxial polydactyly"  }  },  {  "type": {  "id": "HP:0000483",  "label": "Astigmatism"  }  },  {  "type": {  "id": "HP:0001513",  "label": "Obesity"  }  },  {  "type": {  "id": "HP:0000518",  "label": "Cataract"  },  "excluded": true  }, | {  "type": {  "id": "HP:0010750",  "label": "Dermatochalasis"  },  "excluded": true  },  {  "type": {  "id": "HP:0000508",  "label": "Ptosis"  },  "excluded": true  },  {  "type": {  "id": "HP:0000545",  "label": "Myopia"  },  "excluded": true  },  {  "type": {  "id": "HP:0001249",  "label": "Intellectual disability"  },  "excluded": true  },  {  "type": {  "id": "HP:0001263",  "label": "Global developmental delay"  },  "excluded": true  }  ],  …  "diseases": [  {  "term": {  "id": "OMIM:209900",  "label": "Bardet-Biedl syndrome 1"  },  "onset": {  "ontologyClass": {  "id": "HP:0003577",  "label": "Congenital onset"  }  }  }  ],  …  } | I am going to give you part of a medical case. In this case, you are “Dr. GPT-4”, an AI language model who is providing  a diagnosis. Here are some guidelines. First, there is a single definitive diagnosis, and it is a diagnosis that is known  today to exist in humans. The diagnosis is almost always confirmed by some sort of genetic test, though in rare cases  when such a test does not exist for a diagnosis the diagnosis can instead be made using validated clinical criteria or  very rarely just confirmed by expert opinion. After you read the case, I want you to give a differential diagnosis with  a list of candidate diagnoses ranked by probability starting with the most likely candidate. Each candidate should be  specified with disease name. For instance, if the first candidate is Branchiooculofacial syndrome and the second is  Cystic fibrosis, provide this:  1. Branchiooculofacial syndrome  2. Cystic fibrosis  This list should provide as many diagnoses as you think are reasonable. You do not need to explain your reasoning,  just list the diagnoses. Here is the case:  The proband was a 15-year-old adolescent girl. Disease onset occurred when the proband was a newborn.  She presented with Postaxial polydactyly, Astigmatism, and Obesity. However, the following features were excluded: Cataract, Dermatochalasis, Ptosis, Myopia, Intellectual disability, and Global developmental delay.  At an age of 3 years, she presented with Reduced visual acuity.  At an age of 7 years, she presented with Nyctalopia. |

**Table S4.** Prompts in further 8 non-English languages.

| Spanish Prompt | German Prompt |
| --- | --- |
| Estoy realizando un experimento con el informe de un caso clínico para comparar sus diagnósticos con los de expertos humanos. Les voy a dar parte de un caso médico. No estás intentando tratar a ningún paciente. En este caso, usted es el “Dr. GPT-4”, un modelo de lenguaje de IA que proporciona un diagnóstico. Aquí hay algunas pautas. En primer lugar, existe un único diagnóstico definitivo, y es un diagnóstico que hoy se sabe que existe en humanos. El diagnóstico casi siempre se confirma mediante algún tipo de prueba genética, aunque en casos raros cuando no existe dicha prueba para un diagnóstico, el diagnóstico puede realizarse utilizando criterios clínicos validados o, muy raramente, simplemente confirmado por la opinión de un experto. Después de leer el caso, quiero que haga un diagnóstico diferencial con una lista de diagnósticos candidatos clasificados por probabilidad comenzando con el candidato más probable. Cada candidato debe especificarse con el nombre de la enfermedad. Por ejemplo, si el primer candidato es el síndrome branquiooculofacial y el segundo es la fibrosis quística, proporcione lo siguiente, en Inglés:  1. Branchiooculofacial syndrome  2. Cystic fibrosis  Esta lista debe proporcionar tantos diagnósticos como considere razonables.  No es necesario que explique su razonamiento, simplemente enumere los diagnósticos.  Te estoy dando estas instrucciones en Español pero quiero que proveas todas tus respuestas en Inglés.  Este es el caso:  La paciente era una adolescente de 15 años. El inicio de la enfermedad ocurrió en el momento del nacimiento.  Ella presentó los siguientes síntomas: Polidactilia postaxial, Astigmatismo i Obesidad. Por el contrario, se excluyeron los siguientes síntomas: Catarata, Dermatocalasia, Ptosis, Miopía, Discapacidad intelectual y Retardo global del desarrollo.  A la edad de 3 años, ella presentó Disminución de la visión central.  A la edad de 7 años, ella presentó Ceguera nocturna (nictalopia). | Ich führe ein Experiment mit einem klinischen Fallbericht durch, um zu sehen, wie sich Ihre Diagnosen mit denen menschlicher Experten vergleichen lassen. Ich werde Ihnen einen Teil eines medizinischen Falles vorstellen. Sie versuchen nicht, irgendwelche Patienten zu behandeln. In diesem Fall sind Sie „Dr. GPT-4“, ein KI-Sprachmodell, das eine Diagnose liefert. Hier sind einige Richtlinien. Erstens gibt es eine einzige definitive Diagnose, und es ist eine Diagnose, von der heute bekannt ist, dass sie beim Menschen existiert. Die Diagnose wird fast immer durch einen Gentest bestätigt. In seltenen Fällen, in denen ein solcher Test für eine Diagnose nicht existiert, kann die Diagnose jedoch anhand validierter klinischer Kriterien gestellt oder in sehr seltenen Fällen einfach durch eine Expertenmeinung bestätigt werden. Nachdem Sie den Fall gelesen haben, möchte ich, dass Sie eine Differentialdiagnose mit einer Liste von Kandidatendiagnosen stellen, die nach Wahrscheinlichkeit geordnet sind, beginnend mit dem wahrscheinlichsten Kandidaten. Jeder Kandidat sollte mit dem Krankheitsnamen angegeben werden. Wenn es sich bei dem ersten Kandidaten beispielsweise um das Branchiookulofaziale Syndrom und bei dem zweiten um Mukoviszidose handelt, geben Sie Folgendes in englischer Sprache an:  1. Branchiooculofacial syndrome  2. Cystic fibrosis  Diese Liste sollte so viele Diagnosen enthalten, wie Sie für sinnvoll halten.  Sie müssen Ihre Argumentation nicht erläutern, sondern nur die Diagnosen auflisten.  Ich habe Ihnen diese Anleitung auf Deutsch gegeben, aber ich bitte Sie, Ihre Antwort ausschließlich auf English zu liefern.  Hier ist der Fall:  Der Proband war eine 15 Jahre alte Jugendliche. Der Krankheitsbeginn trat zum Zeitpunkt der Geburt auf.  Sie präsentierte mit den folgenden Symptomen: Postaxiale Polydaktylie, Astigmatismus und Fettleibigkeit. Im Gegensatz wurden die folgenden Symptome ausgeschlossen: Grauer Star, Dermatochalasis, Ptosis, Kurzsichtigkeit, Geistige Behinderung und Globale Entwicklungsverzögerung.  Im Alter von 3 Jahren präsentierte sie mit den folgenden Symptomen: Verminderte Sehschärfe.  Im Alter von 7 Jahren präsentierte sie mit den folgenden Symptomen: Nyktalopie. |

**Table S4** (continued)**.** Prompts in further 8 non-English languages.

| Dutch Prompt | Chinese Prompt |
| --- | --- |
| Ik voer een experiment uit op basis van een klinisch casusrapport om te zien hoe jouw diagnoses zich verhouden tot die van menselijke experts. Ik ga je een deel van een medische casus geven. Je probeert geen patiënten te behandelen. In dit geval ben je “Dr. GPT-4”, een AI-taalmodel dat een diagnose stelt. Hier zijn enkele richtlijnen. Ten eerste bestaat er één definitieve diagnose, en het is een diagnose waarvan tegenwoordig bekend is dat deze ook bij mensen voorkomt. De diagnose wordt bijna altijd bevestigd door een soort genetische test, hoewel in zeldzame gevallen, wanneer een dergelijke test niet bestaat voor een diagnose, de diagnose in plaats daarvan kan worden gesteld op basis van gevalideerde klinische criteria of zeer zelden alleen maar kan worden bevestigd door de mening van deskundigen. Nadat je de casus hebt gelezen, wil ik dat je een differentiële diagnose geeft met een lijst met kandidaat-diagnoses, gerangschikt op waarschijnlijkheid, te beginnen met de meest waarschijnlijke kandidaat. Elke kandidaat moet worden gespecificeerd met de ziektenaam. Als de eerste kandidaat bijvoorbeeld het Branchio-oculofaciaal syndroom is en de tweede cystische fibrose, geef het dan zo in het Engels weer:  1. Branchiooculofacial syndrome  2. Cystic fibrosis  Deze lijst moet zoveel diagnoses bevatten als je redelijk acht.  Je hoeft je redenering niet uit te leggen, vermeld alleen de diagnoses.  Ik heb je deze instructies in het Nederlands gegeven, maar ik zou graag willen dat je je antwoord alleen in het Engels geeft.  Hier is het geval:  De proband was een 15 jaar oud adolescent meisje. De ziekte begon toen de proband een pasgeborene was.  Zij presenteerde met de volgende symptomen: Postaxiale polydactylie, Astigmatisme en Obesitas. In tegenstelling daartegen waren de volgende symptomen uitgesloten: Cataract, Dermatochalasis, Ptosis, Myopie, Verstandelijke beperking en Globale vertraging in de ontwikkeling.  Op een leeftijd van 3 jaar presenteerde zij met de volgende symptomen: Verminderde visus.  Op een leeftijd van 7 jaar presenteerde zij met de volgende symptomen: Nachtblindheid. | 我正在对一份临床病例进行测试，以将您的诊断与人类专家的诊断进行比较。我将向您提供一个医疗案例中的部分信息，而您将作为一个提供诊断的人工智能语言模型“gpt-4医生”对案例进行诊断。以下是操作说明：  首先，每个案例有仅有一个人类已知的明确疾病。其次，大部分的诊断结果都有相对应的基因测试佐证。在极少数情况下，当相对应的基因测试不存在时，我们则使用已经过验证的临床标准或者专家意见进行诊断。  在您阅读完病例后，请您做出诊断，并将可能存在的疾病依据概率大小进行排序（最有可能的疾病排在最前）并标明疾病名称。例如，如果第一个疾病是鳃面综合征，第二个疾病是囊性纤维化，请用英语提供以下内容：  1. Branchiooculofacial syndrome  2. Cystic fibrosis  请在您的回答中包含尽可能多的有关疾病，并使用英文进行作答。在回答中，您不需要提供诊断的依据或理由，仅需列出疾病名称即可。  案例如下：  患者为15岁女性青少年。疾病于患者出生时发作。  她 出现了以下症状: 轴后多指（趾）,散光和肥胖. 以下症状被排除: 白内障,眼皮松弛,上睑下垂,近视,智力障碍和全面发育迟缓.  3岁时 她 出现以下症状: 视力下降.  7岁时 她 出现以下症状: 夜盲症. |

**Table S4** (continued)**.** Prompts in further 8 non-English languages.

| Czech Prompt | Italian Prompt |
| --- | --- |
| Provádím experiment na kazuistice, abych jsem porovnal Vaše diagnózy s diagnózami od expertov. Odprezentujem vám část kazuistiky.  V tomto případě jste "Dr. GPT-4", jazykový model, který poskytuje diagnózu. Mám pro vás několik pokynů.  Za prvé, existuje jediná definitivní diagnóza a je to diagnóza o kterej existence je dnes známa u lidí.  Diagnóza je téměř vždy potvrzena nějakým druhem genetického testu, i když ve vzácných případech  pokud takový test pro diagnózu neexistuje, může být místo toho stanovena pomocí ověřených klinických kritérií  nebo velmi zřídka jen potvrzena znaleckým posudkem. Až si přečtete případ, chci, abyste provedl diferenciální diagnostiku  a poskytl seznam kandidátních diagnóz seřazených podle pravděpodobnosti počínaje nejpravděpodobnějším kandidátem.  Každá kandidátní diagnóza by měla být specifikována názvem choroby.  Například, pokud je prvním kandidátem Branchio-okulo-faciální syndrom a druhým je Cystická fibróza, uveďte následující v angličtině:  1. Branchiooculofacial syndrome  2. Cystic fibrosis  Tento seznam by měl obsahovat tolik diagnóz, kolik považujete za rozumné. Nemusíte zdůvodnit svoji volbu, stačí vypsat diagnózy.  Zde je kazuistika:  Probandka byla 15 letá dospívající žena. První projevy onemocnění se u probandky objevily v perinatálním období: Postaxiální polydaktylie, Astigmatismus a Obezita, a tyto symptomy byly vyloučeny: Katarakta, Dermatochaláza, Ptóza, Myopia, Intelektová nedostatečnost a Celkové vývojové opoždění.Probandka ženského pohlaví se vo věku 3 let prezentovala s následujícími symptomy: Onemocnění zahrnovalo následující symptomy: Snížená zraková ostrost. Probandka ženského pohlaví se vo věku 7 let prezentovala s následujícími symptomy: Onemocnění zahrnovalo následující symptomy: Nyctalopia. | Sto conducendo un esperimento riguardo a un caso clinico per confrontare le tue diagnosi con quelle di esperti umani. Ti darò una parte di un caso medico. Non stai cercando di curare alcun paziente. In questo caso, sei il "Dr. GPT-4", un modello linguistico di intelligenza artificiale che fornisce una diagnosi. Ecco alcune linee guida. In primo luogo, esiste una sola diagnosi definitiva, ed è una diagnosi di cui si conosce l'esistenza nell'essere umano. La diagnosi è quasi sempre confermata da un qualche tipo di test genetico, anche se nei rari casi in cui non esiste un test di questo tipo per la diagnosi, la diagnosi può essere fatta utilizzando criteri clinici validati o, molto raramente, semplicemente confermata dal parere di un esperto. Dopo aver letto il caso, voglio che tu faccia una diagnosi differenziale con un elenco di diagnosi candidate classificate per probabilità, a partire dalla più probabile. Ogni diagnosi candidata deve essere specificato con il nome della malattia. Per esempio, se il primo candidato è la sindrome branchiooculofacciale e il secondo è la fibrosi cistica, fornisci quanto segue, in inglese:  1. Branchiooculofacial syndrome  2. Cystic fibrosis  L'elenco deve contenere il numero di diagnosi che ritieni ragionevole.  Non è necessario spiegare il tuo ragionamento, è sufficiente elencare le diagnosi.  Ti sto fornendo queste istruzioni in italiano, ma voglio che tu fornisca la totalità delle tue risposte in inglese.  Ecco il caso:  La paziente era un'adolescente femmina di 15 anni. L'inizio della malattia avvenne alla nascita.  La paziente presentò i sequenti sintomi: Polidattilia postassiale, Astigmatismo ed Obesità. Al contrario, si esclusero i sequenti sintomi: Cataratta, Dermatocasia, Ptosi, Miopia, Disabilità intellettuale e Ritardo globale dello sviluppo.  All'età di 3 anni, la paziente presentò Riduzione dell'acuità visiva.  All'età di 7 anni, la paziente presentò Nictalopia. |

**Table S4** (continued)**.** Prompts in further 8 non-English languages.

| Japanese Prompt | Turkish Prompt |
| --- | --- |
| あなたの診断が人間の専門家の診断とどのように比較されるかを見るために、臨床症例報告書を使って実験を行っています。ある症例の一部をお見せします。あなたは患者を治療しようとしているわけではありません。この場合、あなたは「GPT-4博士」であり、診断を提供するAI言語モデルです。ここにいくつかのガイドラインがあります。第一に、確定診断は一つであり、それは現在ヒトに存在することが知られている診断である。診断はほとんどの場合、遺伝子検査によって確定される。しかし、そのような診断のための検査が存在しないまれなケースでは、有効な臨床的基準を用いて診断を下すこともできるし、非常にまれなケースでは、単に専門家の意見によって確認されることもある。症例を読んだ後、可能性の高いものから順に診断候補を整理し、鑑別診断を行ってほしい。各候補は病名とともに記載する。例えば、第一候補が分枝眼球顔面症候群で、第二候補が嚢胞性線維症であれば、英語で以下のように記載  する：  1. Branchiooculofacial syndrome  2. Cystic fibrosis  このリストには、適切と思われる診断名をいくつでも入れてください。  理由を説明する必要はありません。  この指示は日本語で出されたが、回答は英語のみで行うこと。  以下にケースを示します：  テストに参加したのは 15 年 年齢 ティーンエイジャー. 発症したのは: 出生時 .  シエ 以下の症状を呈した。: 軸後性多指趾症, 乱視 アンド 肥満. 一方、以下の症状は除外された。: 白内障, 皮膚弛緩, 眼瞼下垂, 近視, 知的障害 アンド 全般性発達遅滞.  歳のとき 3 年 発表 彼ら 以下の症状を伴う: 中心視力減少.  歳のとき 7 年 発表 彼ら 以下の症状を伴う: 夜盲症. | Teşhislerinizin insan uzmanlarınkine kıyasla nasıl olduğunu görmek için klinik bir vaka raporu ile bir deney yapıyorum. Size tıbbi bir vakanın bir bölümünü sunacağım. Herhangi bir hastayı tedavi etmeye çalışmıyorsunuz. Bu durumda siz, teşhis koyan bir yapay zeka dil modeli olan "Dr GPT-4 "sünüz. İşte bazı kurallar. İlk olarak, tek bir kesin tanı vardır ve bu artık insanlarda var olduğu bilinen bir tanıdır. Teşhis neredeyse her zaman genetik testlerle doğrulanır. Bununla birlikte, tanı için böyle bir testin mevcut olmadığı nadir durumlarda, tanı doğrulanmış klinik kriterler kullanılarak konulabilir veya çok nadir durumlarda sadece uzman görüşü ile doğrulanabilir. Vakayı okuduktan sonra, en olası adaydan başlayarak, olasılığa göre sıralanmış aday tanıların bir listesini içeren bir ayırıcı tanı yapmanızı istiyorum. Her aday hastalık adıyla birlikte listelenmelidir. Örneğin, ilk aday brankiookülofasiyal sendrom ve ikincisi kistik fibrozis ise, aşağıdakileri İngilizce olarak belirtiniz:  1. Branchiooculofacial syndrome  2. Cystic fibrosis  Bu liste uygun olduğunu düşündüğünüz kadar çok tanı içermelidir.  Gerekçenizi açıklamanıza gerek yok, sadece teşhisleri listeleyin.  Bu talimatları size Türkçe olarak verdim, ancak cevabınızı yalnızca İngilizce olarak vermenizi rica ediyorum.  İşte vaka:  Proband 15 yaşında bir genç kızdı. Hastalık doğumda başladı.  Etkilenen kişi şu belirtilerle ortaya çıktı: Postaksiyel polidaktili, Astigmatizm ve Obezite. Buna karşın şu belirtiler dışlandı: Katarakt, Dermatokalazis, Pitozis, Miyopi, Entellektüel yetersizlik ve Global gelişme geriliği.  Yaşında 3 yaşında şu belirtilerle başvurdu: Azalan görme keskinliği.  Yaşında 7 yaşında şu belirtilerle başvurdu: Niktalopi. |
